# Scaling Safe Resident Driven OPAT bundle in a Lower Middle-Income Country: A Quality Improvement Intervention

**DOI:** 10.64898/2026.07.23.26358342

**Authors:** Amit Mathur, Ravi Kant, VS Pai, Mukesh Bairwa, Darab Singh, Prasan Kumar Panda

**Affiliations:** Department of General Medicine, All India Institute of Medical Sciences, Rishikesh, 249203, India

**Keywords:** Outpatient Parenteral Antimicrobial Therapy (OPAT), Antimicrobial Stewardship, Quality Improvement, Care Bundle, Low- and Middle-Income Countries

## Abstract

**KEY POINTS:** *Question:* To what extent does a standardized quality improvement care bundle improve Outpatient Parenteral Antimicrobial Therapy (OPAT) utilization and clinical outcomes in a resource constrained Indian hospital?

*Findings:* This quality improvement initiative demonstrates that a standardized package of interventions centered on physician education and formal monitoring successfully scaled OPAT utilization to 100% of eligible encounters. The intervention established a highly reliable clinical pathway, maintaining 100% adherence to monitoring indicators without compromising patient safety or increasing hospital readmission rates.

*Meaning:* These results suggest that standardizing the transition to outpatient therapy through a low-cost care bundle can overcome barriers to OPAT utilization in resource-limited settings, achieving universal enrollment and high safety compliance without increasing hospital readmissions.

**Importance:** OPAT is an underutilized strategy in low- and middle-income countries (LMICs). Addressing the knowledge gaps among frontline physicians through structured interventions is vital for optimizing hospital bed utilization and antimicrobial stewardship.

**Objective:** To assess whether a structured multidisciplinary care bundle is associated with improved patient enrollment and clinical care quality in an OPAT program in India.

**Design, Setting, and Participants:** This pre-post quality improvement study was conducted in the Department of General Medicine at a tertiary care referral hospital in Rishikesh, India. Data from patient encounters during a 6-month pre-intervention period (December 1, 2023 to May 31, 2024) were compared with encounters during a 6-month post-implementation period (January 1, 2025 to June 31, 2025). Participants included all postgraduate residents serving as frontline clinical practitioners.

**Interventions:** A structured OPAT bundle comprising a formalized educational curriculum (interactive didactic sessions and bedside practical training), standardized eligibility screening, and a coordinated telephonic monitoring protocol (from June 1, 2024 to December 31, 2024).

**Main Outcomes and Measures:** The primary outcome was the change in the number of eligible patients enrolled in OPAT. Secondary outcomes included clinical process quality indicators (counseling, IV access arrangement, and monitoring compliance), 30-day rehospitalization rates, and therapy-related complications.

**Results:** A total of 20 preintervention patient encounters were compared with 39 postintervention encounters, with similar baseline characteristics between groups (mean age, 37 vs 40 years; male gender, 65% vs 69.2%). Prior to implementation, only 33.3% (20 of 60) of eligible patients received OPAT, whereas 100% (39 of 39) of eligible patients were enrolled following the intervention (p < 0.001). Key clinical processes reached 100% compliance post implementation, including patient counseling (40% vs 100%; p < 0.001), pre-discharge IV access (40% vs 100%; p < 0.001), and daily telephonic monitoring (10% vs 100%; p < 0.001). Safety outcomes remained stable, with no significant differences in readmission rates (5.0% vs 0%; p = 0.34) or drug-related complications.

**Conclusions and Relevance:** This quality improvement study provides evidence that a structured package of OPAT bundle interventions is associated with a transition to universal enrollment of eligible patients and perfect adherence to safety indicators. These results suggest that standardizing the outpatient transition through physician education and coordinated monitoring is a feasible and effective strategy for optimizing hospital resource utilization in LMICs.

## INTRODUCTION

In low- and middle-income countries (LMICs) like India, overstretched healthcare infrastructure, high burden of antimicrobial resistance (AMR), and the high out-of-pocket cost of treatment pose significant challenges to delivering safe and effective antimicrobial therapy. [1] OPAT enables medically stable patients requiring intravenous antimicrobial treatment to complete their therapy outside the hospital, reducing inpatient bed utilization, healthcare costs, and the risk of hospital acquired complications while preserving patient quality of life. [2] Owing to differences in health system infrastructure, availability of multidisciplinary support, monitoring capabilities, and socioeconomic factors, OPAT is widely implemented in middle- and high-income countries (HICs) but remains infrequently practiced in low- and middle-income countries (LMICs).[3] Previous studies from HICs indicate that healthcare professionals are often inadequately trained to deliver OPAT safely and effectively. Knowledge gaps were reported among clinicians in nearly all hospitals offering OPAT in 2022,[4] and both hospital-based and family physicians cited insufficient training on administration devices in 2016. [5] In 2018, infectious disease specialists highlighted a shortage of trained staff to support OPAT services.[6] In India, where OPAT is rarely implemented in resource-limited settings, treating physicians generally lack awareness of the evidence-based guidelines necessary for its safe and effective practice.

To date, no study has systematically evaluated the impact of quality improvement interventions on OPAT practice following the empowerment and targeted training of treating physicians.

The aim of this study was to evaluate whether implementing a structured package of OPAT interventions, also known as OPAT bundle, at a single tertiary-care hospital in India was associated with improvements in clinical care quality.

## METHODS

In this quality improvement study, we performed a pre-post quality improvement project to assess the outcome before and after implementing the OPAT guidelines during a 7-month period from June 1, 2024 - December 31 2024. The study was approved by the Institutional Ethics Committee, All India Institute of Medical Sciences (AIIMS), Rishikesh, India. We adhered to Standards for Quality Improvement Reporting Excellence (SQUIRE) reporting guidelines during the study. [7]

### Setting

This study was conducted in the Department of General Medicine at the AIIMS, Rishikesh, a tertiary-care hospital located in northern India. The inpatient unit of the Department of General Medicine comprises 60 ward beds, distributed equally between two units, of which 24 are designated as high-dependency unit beds. As the majority of patients requiring parenteral antimicrobial therapy are admitted under the Department of General Medicine, this department was selected as the study setting. The selection of a tertiary academic center for this intervention was intentional, aiming to leverage the ‘knowledge cascade’ effect in LMIC settings. As postgraduate residents transition from tertiary training to clinical practice in secondary and primary care settings, they act as vectors for evidence-based OPAT protocols. This strategy ensures the sustainability of the intervention and facilitates the dissemination of specialized antimicrobial stewardship practices to resource-constrained peripheral facilities where formal OPAT infrastructure is currently absent. The well-documented benefits of OPAT, coupled with its poor utilization and the existing knowledge gaps among healthcare professionals, provided the rationale for the design and execution of this study.

### Study Design

All postgraduate residents serving as frontline clinical practitioners were enrolled and provided online consent. None had received prior formal training in OPAT, consistent with prevailing training practices in India.

Baseline knowledge of postgraduate residents was assessed using a multiple-choice questionnaire evaluating OPAT indications, common complications, and implementation principles (eTable 1).

The core intervention strategy was the implementation of the OPAT Bundle, defined as a multidisciplinary care package designed to standardize the transition of care for patients residing across diverse geographic regions, including rural Uttar Pradesh and Uttarakhand of India.

The bundle comprised four core elements:

1. Structured resident physician education,
2. Standardized eligibility checklist for daily patient screening,
3. Pre-discharge vascular access coordination, and
4. Structured tele-monitoring protocol for remote clinical oversight.

The educational component of the bundle included:

1. Interactive didactic sessions,
2. Reinforcement lectures,
3. Bedside practical training, and
4. Dissemination of recorded lecture videos via personal message.

Post-intervention assessment incorporated additional advanced questions to evaluate clinical reasoning, decision-making, and application of OPAT principles in real-world scenarios.

### Data Collection

Data on patients discharged on OPAT during the 6 months preceding program implementation (December 1, 2023 to May 31, 2024) were collected from the Medical Records Department. Pre- and post-training evaluation data for postgraduate residents were obtained using Google Forms. Following the intervention, OPAT discharge data were extracted from the Medical Records Department (January 1, 2025 to June 31, 2025). All data were stored in a secure, password-protected, web-based platform (REDCap [Research Electronic Data Capture]).[8]

### Outcomes

1. Primary process measure:

- OPAT enrollment rate (proportion of eligible patients successfully discharged on OPAT)
2. Secondary process quality indicators:

- Resident training: Change in postgraduate residents’ knowledge scores and clinical reasoning proficiency.
- Care transition: Rates of pre-discharge structured counseling and successful vascular access placement.
- Clinical monitoring: Performance of daily telephonic follow-up and compliance with scheduled Outpatient Department (OPD) review.
3. Clinical outcomes and balancing measures:

- Mean duration of OPAT therapy.
- Treatment completion rates.
- Incidence of drug-related complications and thrombophlebitis.
- All-cause 30-day readmission and mortality.

### Statistical Analysis

Descriptive statistics were used to summarize baseline demographic and clinical characteristics. Continuous variables, such as age, were reported as means with ranges, as the sample size and distribution favored this presentation. Categorical variables, including sex, OPAT setting, administration method, and geographic location, were reported as frequencies and percentages.

To assess the impact of the quality improvement intervention, we compared process and outcome measures between the pre-intervention and post-intervention cohorts. Fishers exact test was employed for all categorical comparisons (including OPAT uptake, clinical process compliance, and safety indicators) because of the small sample sizes and the presence of zero-frequency cells in the post-intervention group (e.g., 100% compliance rates).

For our primary outcome, the proportion of eligible patients receiving OPAT, we calculated the absolute percentage difference and its 95% confidence interval using the Newcombe-Wilson score method to account for proportions reaching 100%.

All p values were two-sided, and a value of p < 0.05 was considered statistically significant. Quality improvement data trends were monitored throughout the study period to ensure process stability and to identify special-cause variation following the implementation of the OPAT bundle.

## RESULTS

### 1. Clinical System Performance and Patient Outcomes

A total of 59 patient encounters were analyzed, comparing 20 patients in the pre-intervention phase with 39 enrolled following the implementation of the OPAT bundle (Table 1). The two cohorts were clinically and demographically comparable; the study population primarily comprised middle-aged males from rural geographic regions. In the majority of cases, antimicrobial therapy was administered in the home setting facilitated by local nursing support.

**Table 1.**
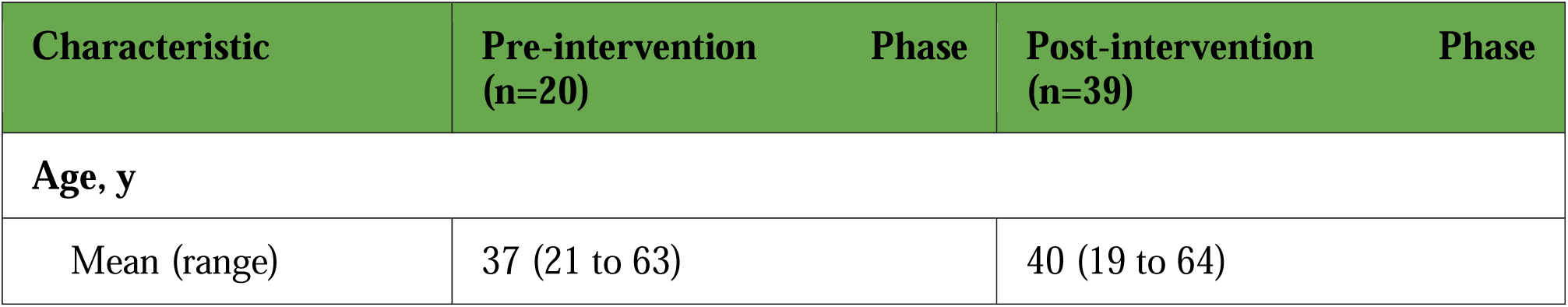
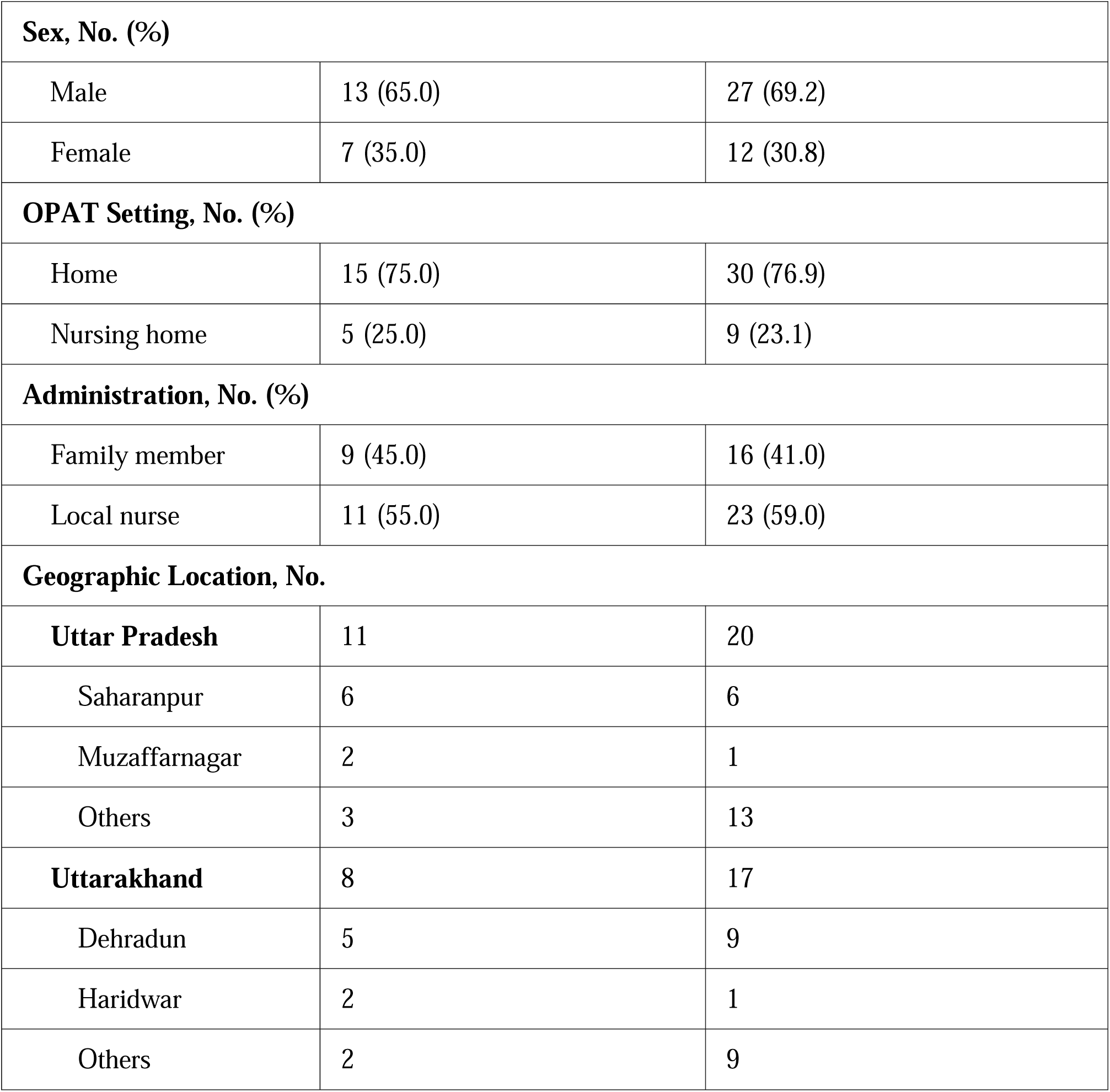
Baseline Demographic and Clinical Characteristics of Patients in the Pre-intervention and Post-intervention OPAT Cohorts.

Prior to program implementation, only 20 of 60 eligible patients (33.3%) received OPAT; following implementation, all 39 eligible patients (100%) received OPAT.

Program implementation was associated with significant improvements across multiple OPAT quality indicators (Table 2). Key clinical processes reached 100% compliance following implementation, including patient counseling, arrangement of intravenous access prior to discharge, and outpatient follow-up compliance (eTable 2).

**Table 2.** Process and Clinical Outcomes Before and After Implementation of the OPAT Bundle.

| **Outcome Measure** | **Pre-intervention**<br>**(n=20)** | **Post-intervention**<br>**(n=39)** | **Difference**<br>**(95% CI)** | **P**<br>**Value** |
| --- | --- | --- | --- | --- |
| **Primary Process Measure** |  |  |  |  |
| OPAT Enrollment, No. (%) | 20/60 (33.3) | 39/39 (100.0) | 66.7(52.8 to 76.4) | <0.001 |
| **Process Quality Indicators** |  |  |  |  |
| Structured counseling, No. (%) | 8 (40.0) | 39 (100.0) | 60.0(38.8 to 74.6) | <0.001 |
| IV access arranged, No. (%) | 8 (40.0) | 39 (100.0) | 60.0(38.8 to 74.6) | <0.001 |
| Daily telephonic follow-up, No. (%) | 2 (10.0) | 39 (100.0) | 90.0(71.1 to 96.7) | <0.001 |
| OPD follow-up compliance, No. (%) | 8 (40.0) | 39 (100.0) | 60.0(38.8 to 74.6) | <0.001 |
| **Clinical and Safety Outcomes** |  |  |  |  |
| Mean OPAT duration, (days) | 14.0 | 8.2 | - | - |
| Premature discontinuation, No. (%) | 2 (10.0) | 0 (0) | -10.0(-30.8 to 4.9) | 0.11 |
| Drug-related complications, No. | 0 | 0 | - | >0.99 |
| Thrombophlebitis, No. (%) | 1 (5.0) | 1 (2.6) | -2.4(-16.5 to 11.9) | >0.99 |
| 30-day Readmission, No. (%) | 1 (5.0) | 0 (0) | -5.0(-23.5 to 9.1) | 0.34 |
| Mortality, No. | 0 | 0 | - | >0.99 |

Monitoring compliance improved significantly, with daily telephonic follow-up reaching 100% saturation in the post-intervention phase. Clinical safety remained robust despite increased utilization; no significant differences were observed in premature OPAT discontinuation or 30-day readmission rates. Adverse events were rare and comparable across both cohorts; no drug-related complications or mortalities were reported, and the incidence of thrombophlebitis was limited to a single patient in each phase.

### 2. Provider Competency and Knowledge Cascade

All residents received training via a multimodal educational program. While didactic sessions and video sharing had the highest attendance, bedside demonstrations provided the most intensive training duration for practical empowerment (eTable 3). However, six residents missed the revision lectures, and eight residents missed the bedside demonstrations.

At baseline, proficiency was suboptimal; over 50% of residents lacked confidence in OPAT practice and were unfamiliar with guidelines, contraindications, and complication monitoring. Notably, over 80% were unaware of necessary follow-up protocols, with a lack of daily monitoring cited as the most frequent clinical error. However, in the post-training phase, more than 90% of residents became aware of OPAT’s purpose, the common infections treated with this approach, and the complications to monitor during follow-up. All trained residents gained confidence in managing complications, understanding the benefits of OPAT, knowing when to change cannulas, and when to stop OPAT.

## DISCUSSION

This study showed that structured OPAT interventions are highly effective in resource-limited settings, where they significantly expanded patient access to therapy. By streamlining the identification and enrollment of eligible candidates, the program achieved universal uptake among eligible patients for OPAT. These findings demonstrate that even in resource-limited settings, standardized clinical pathways can overcome systemic barriers to outpatient antimicrobial delivery without compromising patient safety.[9,10]

Elimination of clinical inertia by standardizing the referral pathway, led to 100% enrollment (up from 33.3%) of OPAT eligible patients in the post implementation phase. The primary driver of the observed improvements was the transformation in physician proficiency. Prior to the intervention, fewer than half of eligible patients were identified for OPAT, reflecting a lack of awareness common in LMICs where formal OPAT training is not part of the standard medical curriculum.[11,12]

Universal compliance with pre-discharge counseling and IV access arrangement suggest that the bundle effectively transitioned these tasks from ad-hoc duties to mandatory steps in the discharge checklist. This ‘bundle’ approach mirrors the success of established OPAT models, where structured coordination and standardized care elements are known to optimize patient outcomes and reduce unplanned readmissions.[11,13]

Our use of daily telephonic monitoring until treatment completion is an intensive oversight model that exceeds the frequency reported in recent HIC cohorts, which typically utilize weekly clinician reviews [14]. This ‘high-touch’ approach is particularly vital in the Indian or LMIC context, as evidenced by recent pilot data from AIIMS Rishikesh [15], which highlights daily monitoring as a primary facilitator for safe OPAT in resource-limited settings. The achievement of nil readmissions in our cohort significantly exceeds the 1.8% readmission rate reported in a recent Indian tertiary care pilot [16], validating that intensive daily telephonic monitoring effectively neutralizes the clinical risks typically associated with the transition from hospital to home care in an LMIC setting. So, in an LMIC setting, daily resident-led telephonic monitoring serves as a high-fidelity, zero-cost safety net that effectively eliminates readmission risk.

Our study observed a notable reduction in mean OPAT duration from 14.0 to 8.2 days following the intervention. This finding aligns with the recent retrospective cohort study by Manders et al. (2025) at Spaarne Gasthuis, which demonstrated that implementing a structured OPAT program with mandatory ID specialist assessment significantly shortened intravenous therapy duration by a mean of 13.97 days (P <0 .001) compared to pre-intervention cohorts. [17]

Clinical safety remained robust throughout the study, with no drug-related complications or mortalities reported. Aside from a single case of thrombophlebitis in each phase (P >0.99), there were no major intravenous access-related complications, and all patients achieved a complete clinical response. These findings align with recent data from a similar Indian tertiary care setting, which demonstrated a high treatment success rate (96.3%) and minimal adverse events, reinforcing that OPAT is a safe and highly effective alternative to inpatient care in the LMIC context when supported by a structured management bundle. [16]

The transition from suboptimal baseline knowledge to near-universal proficiency (>90%) in OPAT guidelines and complication management was a critical driver of the programs success.

By addressing the identified mistakes of the pre-intervention phase, specifically the lack of daily monitoring and inadequate counseling, the multimodal training empowered residents to execute the bundle with 100% compliance. The high levels of post-training confidence in independent practice suggest that structured education is the necessary precursor to system-wide process change.

A unique challenge in Indian tertiary care centers is the high turnover of the resident-led workforce, with new cohorts joining every six months in central institutes and annually in state institutes. In this study, the 6-month intervention window ensured a stable cohort of residents; however, the subsequent departure of four residents upon course completion highlights the necessity of a standardized bundle approach. Because the OPAT bundle is a system-level protocol rather than a person-dependent skill, its quality of care remains intact despite the inevitable rotation of staff. This demonstrates that a structured OPAT approach is highly sustainable in the Indian tertiary care setting, provided the training is integrated into the biannual induction of incoming residents.

## LIMITATION

It is important to acknowledge the limitations of this study.

Firstly, the relatively small sample size of 34 physicians from a single Department of General Medicine may limit the generalizability of the findings to other settings or countries with different healthcare infrastructures and practices. However, since most patients requiring intravenous antimicrobials are typically admitted under General Medicine, the inclusion of 34 treating physicians from this department provides a sufficiently representative sample for evaluating OPAT training outcomes in similar tertiary care settings.

Additionally, the study only involved short-term follow-up, assessing immediate knowledge improvements, which does not address the long-term sustainability of the knowledge gained or its impact on patient outcomes. However, long-term sustainability of knowledge can be supported through a structured peer-learning model, where senior residents mentor juniors, and by regularly circulating recorded video lectures to incoming postgraduate residents.

The use of telephonic monitoring for patient follow-up in the outpatient setting may not have been as effective as face-to-face follow-ups in identifying and managing complications. To address this, developing a specialized support system, such as a telemedicine society or platform, can enhance remote monitoring. Incorporating video calls for follow-ups would allow healthcare providers to visually assess patients, identify complications more effectively, and offer real-time guidance. Such advancements would improve the overall effectiveness of outpatient monitoring while reducing the dependency on in-person visits. Another limitation is the applicability of this training intervention in peripheral healthcare settings such as primary and secondary level care, where there is no resident system. However, the transition of trained residents from tertiary to these peripheral facilities will address this issue for the long run and they can act as mentors to local healthcare workers, ensuring the safe and effective implementation of OPAT at the grassroots level. It was challenging to ensure full compliance with the recorded video lecture, as there was no objective method to track how much of the content was actually watched by each resident. This may have affected the uniformity of training received across participants. Furthermore, the study did not address the underlying infrastructural and resource limitations that continue to pose challenges for OPAT implementation in resource-constrained settings.

## CONCLUSION

In this resource-limited LMIC, a standardized OPAT bundle achieved universal uptake, reduced intravenous treatment duration, and maintained excellent safety. Protocolized referral, mandatory discharge processes, and structured physician training eliminated clinical inertia and care variability, while daily resident-led telephonic monitoring provided a low-cost, high-reliability safeguard with zero readmissions. By embedding OPAT bundle as a system-level process rather than an individual-dependent skill, the intervention proved durable despite workforce turnover. Despite single-center and short-term limitations, this bundled approach represents a feasible, scalable, and safe model for expanding OPAT access in LMICs.

## DATA SHARING STATEMENT

Everything is provided in the manuscript, if any other specific things required, they will be provided with correspondence to author.

## Data Availability

All data produced in the present study are available upon reasonable request to the authors

---

**eTable 1.**
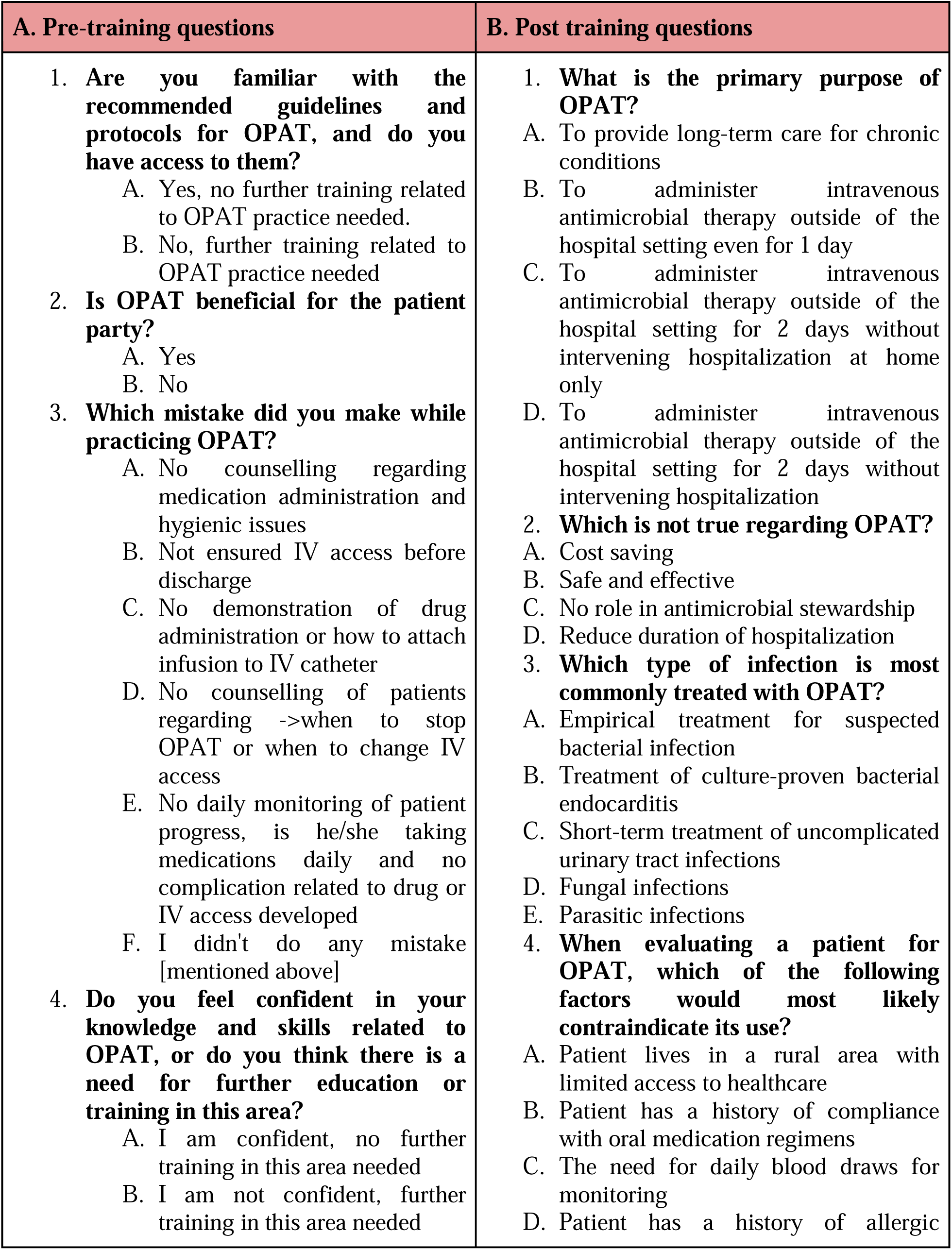
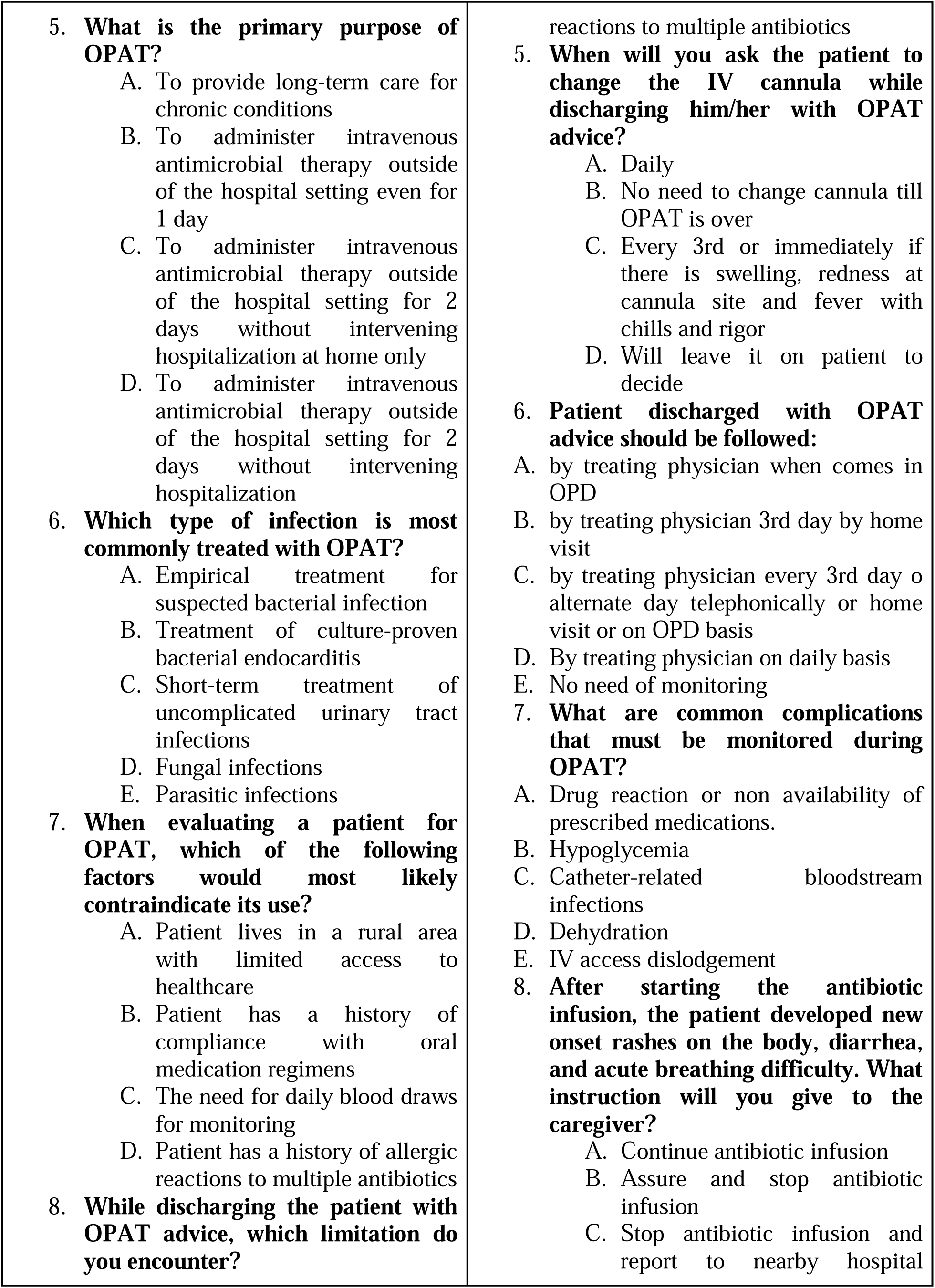
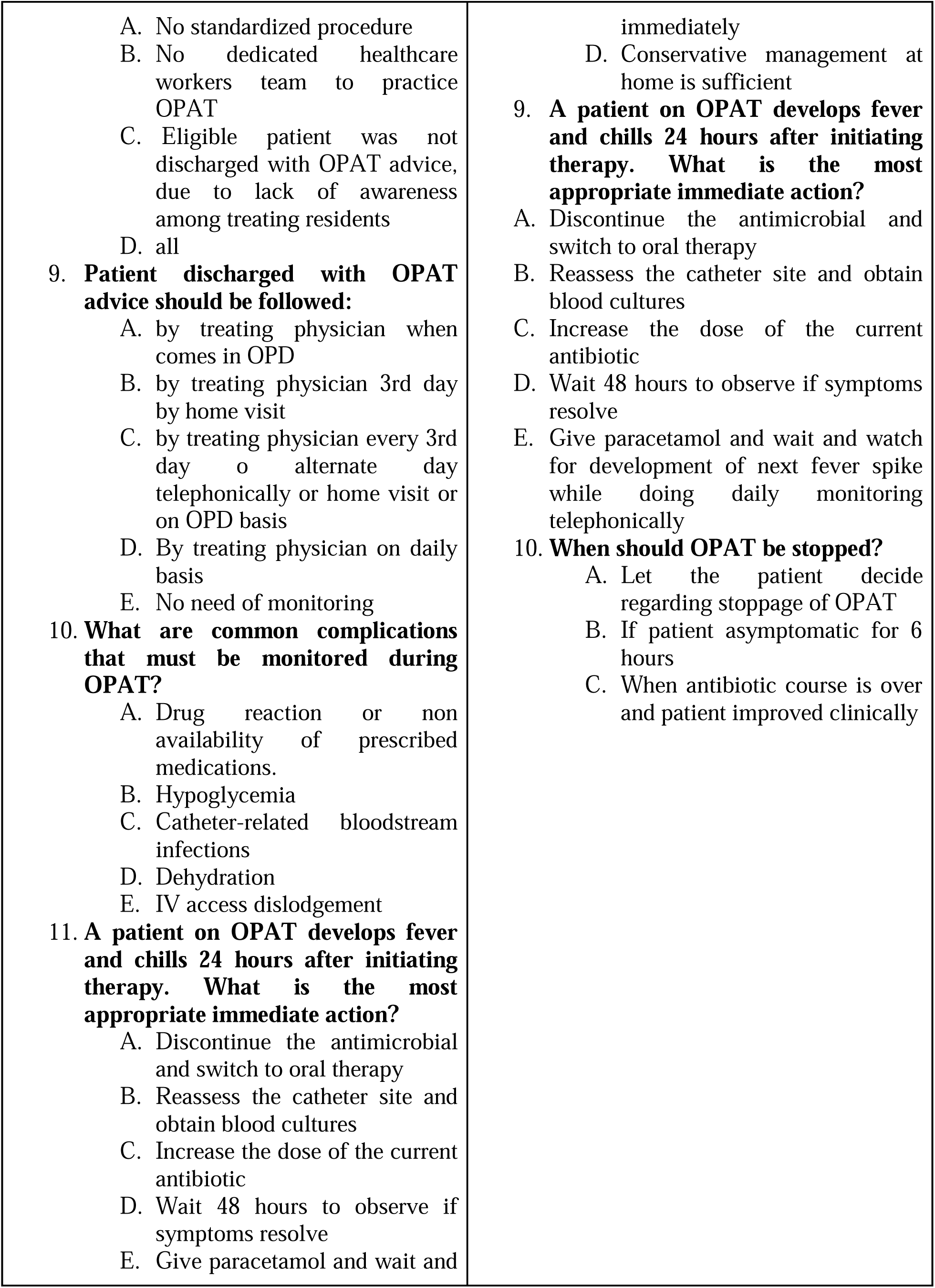
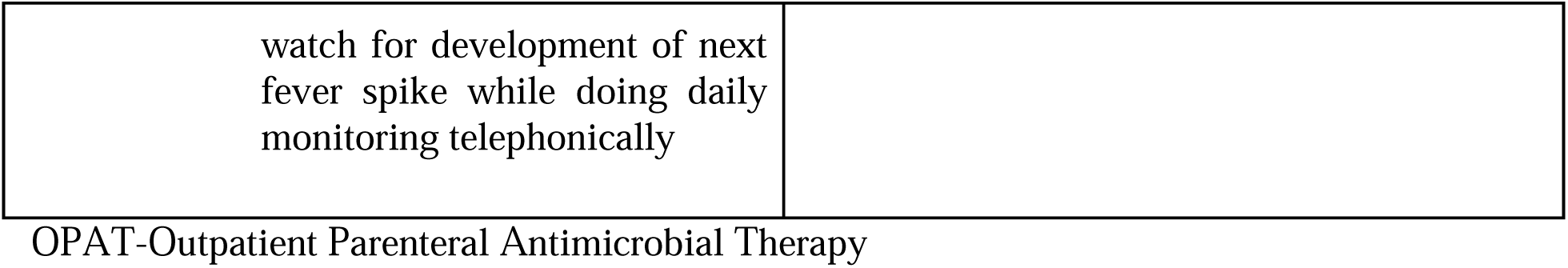
A. Pre-training questions B. Post training questions.

| A. Pre-training questions | B. Post training questions |
| --- | --- |
| 1. **Are you familiar with the recommended guidelines and protocols for OPAT, and do you have access to them?**<br>A. Yes, no further training related to OPAT practice needed.<br>B. No, further training related to OPAT practice needed<br>2. **Is OPAT beneficial for the patient party?**<br>A. Yes<br>B. No<br>3. **Which mistake did you make while practicing OPAT?**<br>A. No counselling regarding medication administration and hygienic issues<br>B. Not ensured IV access before discharge<br>C. No demonstration of drug administration or how to attach infusion to IV catheter<br>D. No counselling of patients regarding ->when to stop OPAT or when to change IV access<br>E. No daily monitoring of patient progress, is he/she taking medications daily and no complication related to drug or IV access developed<br>F. I didn't do any mistake [mentioned above]<br>4. **Do you feel confident in your knowledge and skills related to OPAT, or do you think there is a need for further education or training in this area?**<br>A. I am confident, no further training in this area needed<br>B. I am not confident, further training in this area needed | 1. **What is the primary purpose of OPAT?**<br>A. To provide long-term care for chronic conditions<br>B. To administer intravenous antimicrobial therapy outside of the hospital setting even for 1 day<br>C. To administer intravenous antimicrobial therapy outside of the hospital setting for 2 days without intervening hospitalization at home only<br>D. To administer intravenous antimicrobial therapy outside of the hospital setting for 2 days without intervening hospitalization<br>2. **Which is not true regarding OPAT?**<br>A. Cost saving<br>B. Safe and effective<br>C. No role in antimicrobial stewardship<br>D. Reduce duration of hospitalization<br>3. **Which type of infection is most commonly treated with OPAT?**<br>A. Empirical treatment for suspected bacterial infection<br>B. Treatment of culture-proven bacterial endocarditis<br>C. Short-term treatment of uncomplicated urinary tract infections<br>D. Fungal infections<br>E. Parasitic infections<br>4. **When evaluating a patient for OPAT, which of the following factors would most likely contraindicate its use?**<br>A. Patient lives in a rural area with limited access to healthcare<br>B. Patient has a history of compliance with oral medication regimens<br>C. The need for daily blood draws for monitoring<br>D. Patient has a history of allergic |
| 5. **What is the primary purpose of OPAT?**<br>A. To provide long-term care for chronic conditions<br>B. To administer intravenous antimicrobial therapy outside of the hospital setting even for 1 day<br>C. To administer intravenous antimicrobial therapy outside of the hospital setting for 2 days without intervening hospitalization at home only<br>D. To administer intravenous antimicrobial therapy outside of the hospital setting for 2 days without intervening hospitalization<br>6. **Which type of infection is most commonly treated with OPAT?**<br>A. Empirical treatment for suspected bacterial infection<br>B. Treatment of culture-proven bacterial endocarditis<br>C. Short-term treatment of uncomplicated urinary tract infections<br>D. Fungal infections<br>E. Parasitic infections<br>7. **When evaluating a patient for OPAT, which of the following factors would most likely contraindicate its use?**<br>A. Patient lives in a rural area with limited access to healthcare<br>B. Patient has a history of compliance with oral medication regimens<br>C. The need for daily blood draws for monitoring<br>D. Patient has a history of allergic reactions to multiple antibiotics<br>8. **While discharging the patient with OPAT advice, which limitation do you encounter?** | reactions to multiple antibiotics<br>5. **When will you ask the patient to change the IV cannula while discharging him/her with OPAT advice?**<br>A. Daily<br>B. No need to change cannula till OPAT is over<br>C. Every 3rd or immediately if there is swelling, redness at cannula site and fever with chills and rigor<br>D. Will leave it on patient to decide<br>6. **Patient discharged with OPAT advice should be followed:**<br>A. by treating physician when comes in OPD<br>B. by treating physician 3rd day by home visit<br>C. by treating physician every 3rd day or alternate day telephonically or home visit or on OPD basis<br>D. By treating physician on daily basis<br>E. No need of monitoring<br>7. **What are common complications that must be monitored during OPAT?**<br>A. Drug reaction or non availability of prescribed medications.<br>B. Hypoglycemia<br>C. Catheter-related bloodstream infections<br>D. Dehydration<br>E. IV access dislodgement<br>8. **After starting the antibiotic infusion, the patient developed new onset rashes on the body, diarrhea, and acute breathing difficulty. What instruction will you give to the caregiver?**<br>A. Continue antibiotic infusion<br>B. Assure and stop antibiotic infusion<br>C. Stop antibiotic infusion and report to nearby hospital |
| A. No standardized procedure<br>B. No dedicated healthcare workers team to practice OPAT<br>C. Eligible patient was not discharged with OPAT advice, due to lack of awareness among treating residents<br>D. all<br>9. **Patient discharged with OPAT advice should be followed:**<br>A. by treating physician when comes in OPD<br>B. by treating physician 3rd day by home visit<br>C. by treating physician every 3rd day o alternate day telephonically or home visit or on OPD basis<br>D. By treating physician on daily basis<br>E. No need of monitoring<br>10. **What are common complications that must be monitored during OPAT?**<br>A. Drug reaction or non availability of prescribed medications.<br>B. Hypoglycemia<br>C. Catheter-related bloodstream infections<br>D. Dehydration<br>E. IV access dislodgement<br>11. **A patient on OPAT develops fever and chills 24 hours after initiating therapy. What is the most appropriate immediate action?**<br>A. Discontinue the antimicrobial and switch to oral therapy<br>B. Reassess the catheter site and obtain blood cultures<br>C. Increase the dose of the current antibiotic<br>D. Wait 48 hours to observe if symptoms resolve<br>E. Give paracetamol and wait and | immediately<br>D. Conservative management at home is sufficient<br>9. **A patient on OPAT develops fever and chills 24 hours after initiating therapy. What is the most appropriate immediate action?**<br>A. Discontinue the antimicrobial and switch to oral therapy<br>B. Reassess the catheter site and obtain blood cultures<br>C. Increase the dose of the current antibiotic<br>D. Wait 48 hours to observe if symptoms resolve<br>E. Give paracetamol and wait and watch for development of next fever spike while doing daily monitoring telephonically<br>10. **When should OPAT be stopped?**<br>A. Let the patient decide regarding stoppage of OPAT<br>B. If patient asymptomatic for 6 hours<br>C. When antibiotic course is over and patient improved clinically |
| watch for development of next fever spike while doing daily monitoring telephonically |  |

OPAT-Outpatient Parenteral Antimicrobial Therapy

**eTable 2.**
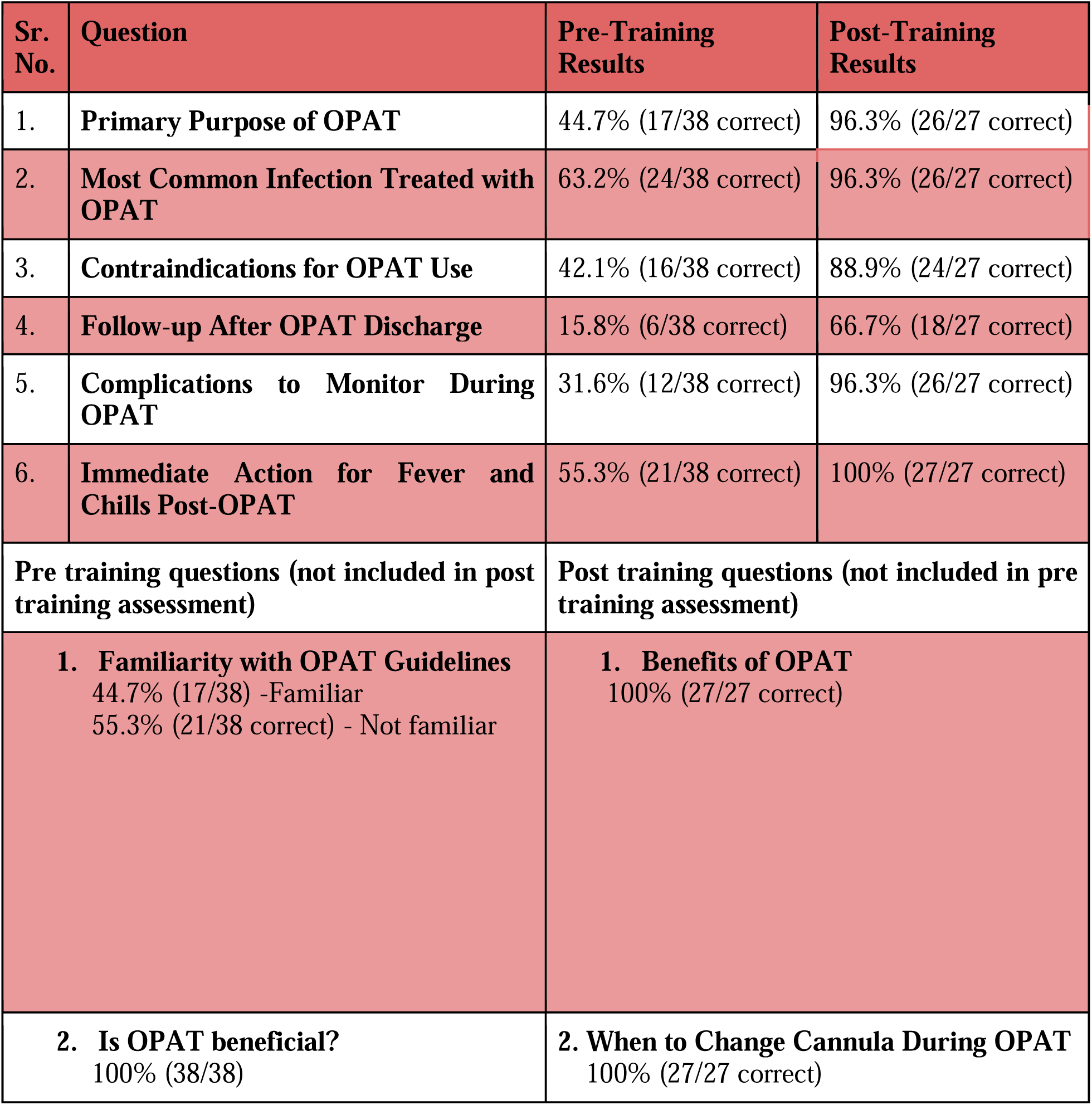
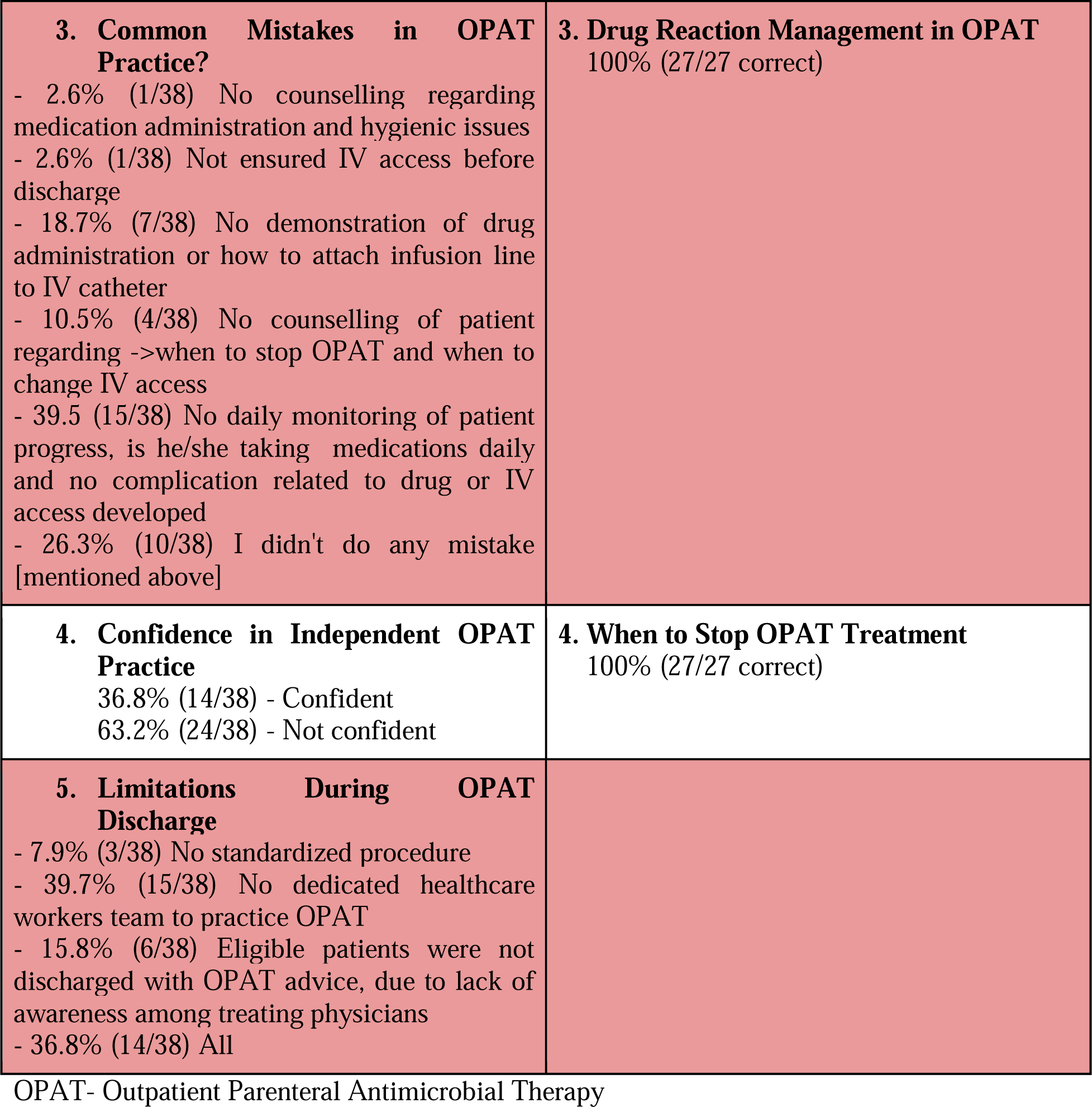
Process Measure: Comparison of Baseline and Post-Intervention OPAT Knowledge and Confidence Among Frontline Physicians.

| **Sr. No.** | **Question** | **Pre-Training Results** | **Post-Training Results** |
| --- | --- | --- | --- |
| 1. | **Primary Purpose of OPAT** | 44.7% (17/38 correct) | 96.3% (26/27 correct) |
| 2. | **Most Common Infection Treated with OPAT** | 63.2% (24/38 correct) | 96.3% (26/27 correct) |
| 3. | **Contraindications for OPAT Use** | 42.1% (16/38 correct) | 88.9% (24/27 correct) |
| 4. | **Follow-up After OPAT Discharge** | 15.8% (6/38 correct) | 66.7% (18/27 correct) |
| 5. | **Complications to Monitor During OPAT** | 31.6% (12/38 correct) | 96.3% (26/27 correct) |
| 6. | **Immediate Action for Fever and Chills Post-OPAT** | 55.3% (21/38 correct) | 100% (27/27 correct) |
| **Pre training questions (not included in post training assessment)** |  | **Post training questions (not included in pre training assessment)** |  |
| **1. Familiarity with OPAT Guidelines**<br>44.7% (17/38) -Familiar<br>55.3% (21/38 correct) - Not familiar |  | **1. Benefits of OPAT**<br>100% (27/27 correct) |  |
| **2. Is OPAT beneficial?**<br>100% (38/38) |  | **2. When to Change Cannula During OPAT**<br>100% (27/27 correct) |  |

|  |  |
| --- | --- |
| **3. Common Mistakes in OPAT Practice?**<br>- 2.6% (1/38) No counselling regarding medication administration and hygienic issues<br>- 2.6% (1/38) Not ensured IV access before discharge<br>- 18.7% (7/38) No demonstration of drug administration or how to attach infusion line to IV catheter<br>- 10.5% (4/38) No counselling of patient regarding ->when to stop OPAT and when to change IV access<br>- 39.5 (15/38) No daily monitoring of patient progress, is he/she taking medications daily and no complication related to drug or IV access developed<br>- 26.3% (10/38) I didn't do any mistake [mentioned above] | **3. Drug Reaction Management in OPAT**<br>100% (27/27 correct) |
| **4. Confidence in Independent OPAT Practice**<br>36.8% (14/38) - Confident<br>63.2% (24/38) - Not confident | **4. When to Stop OPAT Treatment**<br>100% (27/27 correct) |
| **5. Limitations During OPAT Discharge**<br>- 7.9% (3/38) No standardized procedure<br>- 39.7% (15/38) No dedicated healthcare workers team to practice OPAT<br>- 15.8% (6/38) Eligible patients were not discharged with OPAT advice, due to lack of awareness among treating physicians<br>- 36.8% (14/38) All |  |

OPAT- Outpatient Parenteral Antimicrobial Therapy

**eTable 3.** Overview of Training Methods Used for empowerment of residents regarding OPAT.

OPAT- Outpatient Parenteral Antimicrobial Therapy **eTable 3.** Overview of Training Methods Used for empowerment of residents regarding OPAT
| Training Method | Session (n) | Mean duration (Min)+SD | Resident attended |
| --- | --- | --- | --- |
| Interactive lectures | 6 | 38.3 +- 4.71 | 34 |
| Revision lectures | 5 | 33.6 +- 3.85 | 28 |
| Bedside practical demonstration of OPAT practice | 2 | 52.5 +- 2.5 | 26 |
| Recorded lecture video sharing | 35 | 9.27 | 34 |

OPAT-Outpatient Parenteral Antimicrobial Therapy

